# Efficacy of intravenous paracetamol and mannitol in managing chronic post-thoracotomy pain in patients with lung cancer: Study protocol for a single center prospective randomized double-blind controlled trial

**DOI:** 10.1101/2024.08.27.24312675

**Authors:** Xing Lu, Junhui Zhou, Xi Li, Jie Gao, Siqing Liu, Wei Zhong, Gaoyuan Xi, Yingchun Guo, Hongdang Xu

**Author notes:** Corresponding author (HX). These authors contributed equally to this work and are co-first authors (Xing Lu, Junhui Zhou).

## Abstract

Chronic post-thoracotomy pain is a common complication that affects 20% to 60% of patients who undergo surgery for lung cancer. The persistent pain affects quality of life and satisfaction with surgery. Intravenous paracetamol and mannitol, known to have analgesic and antipyretic properties, may help relieve moderate-to-severe post-operative pain. This trial aims to assess their effectiveness and safety in managing chronic post-thoracotomy pain in patients with lung cancer. This prospective double-blind randomized controlled clinical trial will be conducted at a single center. A total of 856 patients who will undergo thoracoscopic radical surgery for lung cancer will be enrolled and randomly assigned to test (intravenous paracetamol and mannitol) and control (intravenous normal saline) groups in a 1:1 ratio (428 patients in each group). Efficacy will be evaluated in terms of the incidence of chronic post-thoracotomy pain at 3 months (primary outcome). Secondary outcomes will include the dosage of propofol and remifentanil, numerical rating scale pain scores, number of times the patient-controlled intravenous analgesia button is pressed, occurrence of post-operative nausea and vomiting and respiratory depression, time to first flatus and ambulation after surgery, length of hospital stay, surgeon and patient satisfaction, and incidence of chronic post-thoracotomy pain at 6 and 12 months after surgery. Quality of daily life will be evaluated at 3, 6, and 12 months after surgery. Intention-to-treat analysis will also be conducted. The research protocol has been reviewed and approved by the Medical Thesis Committee of Henan Provincial Chest Hospital and Chest Hospital of Zhengzhou University on April 29, 2023 (reference: [2023] approval number: [04-06]). The results of this trial will be communicated to the participants and subsequently submitted for publication in peer-reviewed journals for wider dissemination. The study has been registered in the Chinese Clinical Trial Registry on June 27, 2023 (registration number: ChiCTR2300072869, available at https://www.chictr.org.cn/). The protocol version number is Version 1.1, dated August 20, 2023.

## Introduction

Chronic post-surgical pain (CPSP) is common following surgery at different sites and may develop after abdominal, orthopedic, and cardiothoracic procedures. The surgical site and methods are the primary factors that influence the incidence of CPSP [1]. Thoracotomy is a procedure that allows access to the thoracic cavity. As this requires dissection through multiple layers of skin, fat, and muscles, patients experience acute or chronic pain after surgery; notably, thoracic surgery is associated with the highest incidence of chronic pain [2]. Chronic post-thoracotomy pain (CPTP) is a persistent and debilitating condition that affects a substantial number of patients who undergo surgery for lung cancer. This long-term neuropathic pain mainly occurs around the incision site [3]. As this is a type of CPSP, it is defined by persistence for at least 3 months after the procedure [4]. CPTP can significantly impact quality of life in affected patients, and thereby lead to physical and psychological distress; it also increases subsequent medical expenses and social costs. However, an incomplete understanding of its pathophysiology, a lack of personalized therapeutic strategies, absence of standardized assessment criteria, inadequate long-term follow-up, and deficiencies in interdisciplinary teamwork create substantial challenges to the effective management of CPTP. Addressing these issues requires further research, clinical evaluation, and policy support. This may help improve management of the condition and patient quality of life.

Trauma during thoracic surgery typically involves injuries to the costal cartilage, costovertebral joints, muscles, pleura (due to traction), intercostal nerves, and lung parenchyma; thoracic drainage tubes are also a source of irritation. In conjunction, these injuries may contribute to the development of CPTP [5]. Tissue and nerve damage can result in inflammation and edema, and directly cause symptoms of nerve compression. In addition, the release of inflammatory mediators such as interleukin-6, interleukin-8, tumor necrosis factor-α, and substance P may lower the pain threshold by enhancing pain receptor activity. This cascade ultimately results in heightened sensitization of pain centers in the spinal dorsal horn neurons and cerebral cortex. Current understanding therefore supports the notion that the pathogenesis of CPTP involves a combination of somatic, visceral, and neuropathic pain, that is often accompanied by central nervous system sensitization [6].

The presence of intense acute pain following surgery has been identified as one of the predictive risk factors for the development of CPSP [7]. Anesthesiologists therefore use a variety of analgesic techniques to alleviate early post-operative pain in patients undergoing thoracic surgery. Commonly utilized strategies include administration of thoracic epidural analgesia, intravenous non-steroidal analgesics or opioids, and ultrasound-guided nerve blocks (such as paravertebral, intercostal nerve, and erector spinae and serratus anterior plane blocks, among others) [8]. Among these approaches, ultrasound-guided nerve blocks have gained particular popularity for post-operative pain management. This may be attributed to their efficacy in providing analgesia and reducing the need for opioids, both of which potentially decrease opioid-related adverse effects such as post-operative nausea and vomiting [9]. However, nerve block procedures require considerable skill and are associated with risks of complications such as pneumothorax, hematoma, and block failure. Anesthesiologists need to possess a thorough understanding of the position of anatomical structures to address these challenges; this can be particularly demanding for less experienced practitioners.

Paracetamol (acetaminophen) is a 4-p-aminophenol derivative and a type of acetanilide central antipyretic analgesic. Unlike non-steroidal anti-inflammatory drugs, it does not cause damage to the gastric mucosa (that causes indigestion) or kidney. In addition, it does not cause the unpleasant side-effects associated with opioids such as respiratory depression, nausea, vomiting, constipation, or urinary retention [10]. Owing to the lesser potential for adverse reactions and high safety, it is often the first choice for multimodal analgesia after surgery. As per the American Society of Anesthesiologists Task Force on Acute Pain Management [11], post-operative patients should receive non-opioid analgesics as first-line drugs in the absence of any contraindications; opioids are only recommended as auxiliary drugs.

The intravenous route is ideal for administration of paracetamol immediately after surgery, as patients are unable to take oral drugs and/or need rapid pain relief [12]. Previous studies have confirmed that paracetamol injections offer reliable and optimal analgesic effect, while avoiding the adverse reactions of opioids [13]. A systematic review and meta-analysis comprising 5 randomized controlled trials (including 2635 patients) evaluated the impact of intravenous acetaminophen on post-operative pain in those undergoing brain surgery. The results showed that acetaminophen significantly decreased rescue analgesic consumption, total analgesic use, intensive care unit stay, and visual analog scale scores, and increased patient satisfaction. Preoperative administration of intravenous acetaminophen was therefore found to effectively alleviate post-operative pain, reduce analgesic requirements, and improve outcomes in patients undergoing brain surgery [14]. Another study on patients who underwent partial mastectomy showed that the analgesic effect of intravenous acetaminophen was comparable to that of flurbiprofen axetil. However, it was more effective in controlling movement-induced pain at 3 h after surgery, and demonstrated a lower incidence of adverse reactions [15]. Yet another study on patients who underwent video-assisted thoracic surgery demonstrated that the post-operative pain control offered by intravenous ketorolac and acetaminophen was comparable to that of morphine. Notably, the combination offered superior pain control compared to morphine within 2 hours of surgery, and was not associated with any significant adverse reactions [16]. Nevertheless, there is a paucity of long-term follow-up data regarding the use of intravenous acetaminophen. The available studies mainly focus on short-term outcomes, and do not provide crucial long-term follow-up data that are necessary for assessing the sustained effectiveness and safety profile of this drug.

In China, intravenous paracetamol and mannitol was introduced as a novel analgesic combination in 2022. The primary component is the 4-aminophenol derivative, which is known for its analgesic and antipyretic properties [17] and effectively complements opioid analgesics in managing moderate-to-severe post-operative pain in adult patients. The combined analgesic and anti-inflammatory effects of intravenous paracetamol and mannitol show promise in preventing the onset and reducing the progression of CPTP.

The aim of this trial is to evaluate and compare the efficacy and safety of intravenous paracetamol and mannitol versus normal saline in alleviating CPTP in patients with lung cancer. The findings of this trial may help provide crucial clinical evidence for diversifying management strategies in CPTP; this may in turn improve treatment outcomes and quality of life.

## Materials and methods

### Study design

This single-center prospective randomized double-blind controlled trial will adopt a parallel group design. It will aim to investigate the efficacy of intravenous paracetamol and mannitol in managing CPTP among patients who undergo surgery for lung cancer. To maintain the double-blind design, neither the researchers nor the participants will be aware of the assigned study group. This will reduce the impact of subjective bias and ensure objectivity of the results. This trial was registered in the Chinese Clinical Trial Registry on June 27, 2023, with a registration number of ChiCTR2300072869.

### Type of trial

In this parallel-group design trial, participants will be randomly assigned to one of two groups: test group (receiving intravenous paracetamol and mannitol) and control group (receiving normal saline).

### Allocation ratio

Participants will be allocated to either group in a ratio of 1:1; this implies that an equal number of participants will be allocated to each group.

### Framework

This superiority trial aims to demonstrate superiority of the intervention agent (intravenous paracetamol and mannitol) over the control agent (normal saline) in achieving control of CPTP in patients with lung cancer. The study will assess the effectiveness of the intervention in reducing pain intensity and improving overall quality of life (compared to the control). The investigators will strictly adhere to the Standard Protocol Items: Recommendations for Interventional Trials (SPIRIT) guidelines [18] (available for reference on the official website: https://www.spirit-statement.org). The Checklist. SPIRIT checklist can be found in S1 File.

### Participants

This trial will be implemented in an academic hospital, and the data will be collected in China. The eligibility criteria were determined by a researcher who will not be associated with the study; the selected criteria are based on patient parameters at admission. Patients enrolled in this study or their authorized representatives will be required to sign an informed consent form by a trained researcher prior to initiation of treatment. The template for the informed consent form can be found in S2 File.

Patients will be eligible for inclusion if they: 1) are aged between 18 and 64 years; 2) are diagnosed with lung cancer before surgery (or during surgery via rapid frozen pathological examination) and are scheduled to undergo thoracoscopic surgery (such as lobectomy, wedge resection, or anatomic segmentectomy); 3) have American Society of Anesthesiologists grade I–III physical status; and 4) are able to understand the purpose of the study, agree to participate, and provide written informed consent.

Patients will be excluded from this trial if they: 1) only receive conservative treatment without surgery during hospitalization; 2) have any mental illness or cognitive impairment and are unable to cooperate with the study requirements; 3) are diagnosed with severe coagulation dysfunction; 4) have any serious infections; 5) are dependent on drugs or have a history of long-term use of psychotropic drugs; 6) have a history of long-term use of steroids or hormonal agents; 7) have Gilbert syndrome; 8) have an inadequate hepatic glutathione reserve due to chronic malnutrition, anorexia, overeating, or cachexia; 9) suffer from glucose-6-phosphatase dehydrogenase deficiency, which may lead to hemolytic anemia; and 10) are allergic to acetaminophen, mannitol, or other anesthetic drugs.

Trial participation may be terminated if: 1) patients or their families actively request for withdrawal from the study during the study period; 2) temporary thoracotomy closure is necessary during the operation; 3) patients develop severe heart, brain, liver, or kidney dysfunction after surgery; 4) any of the following events occur: failed post-operative recovery, automatic discharge, or perioperative death; and 5) patients or their families cannot be contacted on three or more occasions during the follow-up period.

### Randomization

A computer-generated randomization table will be used to create group random sequences. The random sequences will be generated by independent researchers and stored in secure electronic files to ensure confidentiality of grouping information. The generation of random sequences will ensure that participant grouping is random and unpredictable.

Factors for stratification will include age, gender, and pain severity. To reduce predictability of the random sequence, details of any planned restrictions (such as blocking) will be outlined in a separate document that cannot be accessed by those involved in participant enrollment or intervention assignment.

The specific steps will be as follows: a) a computer will be used to generate random numbers in advance; these will be placed in sequentially numbered, sealed, opaque envelopes which will be managed by designated personnel, b) the patients who meet the inclusion criteria will be numbered according to the order of enrollment, c) random numbers will be matched with those of the patient to obtain the allocated group, d) the results of randomized grouping will be handed over to an anesthesia nurse, who will provide either acetaminophen and mannitol for intravenous administration or normal saline. The appearance, packaging, and labeling of these two drugs will be identical. The syringes will only be labeled as “test drugs” to ensure that the patients, anesthesiologists, and outcome assessors are unaware of the allocated group. The anesthesiologist will administer the drugs in sequential order. The random coding table for the clinical trial (known as a blind bottom) will be kept at the Clinical Trial Center of the Henan Provincial Chest Hospital & Chest Hospital of Zhengzhou University.

Sequentially numbered opaque sealed envelopes will be used to conceal the sequence, and each envelope will contain a unique allocation code corresponding to either the paracetamol and mannitol or normal saline groups. These envelopes will be opened sequentially by a designated research pharmacist only at the time of participant enrollment and assignment to ensure allocation concealment. The allocation codes will not be disclosed to the investigators, participants, or outcome assessors until study completion. This will prevent disclosure of grouping information during the allocation process, and ensure appropriate randomization and blinding.

### Blinding

All anesthesiologists, anesthesia nurses, beneficiaries, and personnel involved in data collection and analysis will be blinded. An anesthesiologist who will be blinded to patient grouping will conduct the follow-up assessments for acute and chronic post-operative pain. The Data Monitoring Committee will establish and supervise emergency unblinding procedures to ensure study integrity.

### Interventions

#### Anesthetic management

All patients will receive anesthesia as per standardized protocols. Routine monitoring of the blood pressure, electrocardiogram, and oxygen saturation will be performed after the patient enters the operating room. Venous access will be established and invasive arterial blood pressure monitoring will be performed. All patients will be administered combined intravenous and inhalational anesthesia. The following drugs will be administered intravenously during induction: midazolam (0.04 mg/kg), etomidate (0.3 mg/kg), sufentanil (0.4 µg/kg), and rocuronium (0.8 mg/kg). Double-lumen endotracheal intubation will then be performed under visual guidance for mechanical ventilation. Following successful tracheal intubation, right internal jugular vein cannulation will be performed under ultrasound guidance. The bispectral index will be monitored continuously during surgery to ensure that the values are maintained within a range of 40–60. Inhalational sevoflurane (1%) will be administered along with propofol (4–8 mg·kg^−1^·h^−1^ by continuous infusion), remifentanil (0.05–0.2 μg·kg^−1^·min^−1^), and rocuronium (0.3–0.6 mg·kg^−1^·h^−1^). The tidal volume, respiratory rate, positive end-expiratory pressure, and fraction of inspired oxygen will be adjusted as appropriate. Balanced crystalloid fluids will be continuously infused at a rate of 3 ml·kg−1·h^−1^ to maintain urine outputs of > 0.5 ml·kg^−1^·h^−1^. Rocuronium will be discontinued 30 min before the end of surgery, and sugammadex will be used for reversing neuromuscular blockade. The double-lumen endotracheal tube will then be removed, and a patient-controlled intravenous analgesia (PCIA) pump will be connected to the intravenous access before transferring the patient to the post-anesthesia care unit (PACU). All patients will undergo thoracoscopic radical resection of the malignant lung lesion, and the procedures will be performed by the same group of surgeons.

#### Study interventions

Patients in the test group will be administered intravenous paracetamol (500 mg) and mannitol 15 min before the end of the operation; they will also be administered intravenous paracetamol (500 mg) and mannitol on post-operative days 1 and 2, respectively. Patients in the control group will be administered intravenous normal saline (50 mL) at the same time points.

#### Pain management

Patients in both groups will be administered intravenous oxycodone (5 mg) 15 min before the end of surgery and PCIA immediately after surgery. The medications in the analgesic pump will include sufentanil (2.5 μg/kg) and tropisetron (10 mg), and will be diluted with normal saline to achieve a total volume of 100 mL. The following analgesic pump parameters will be used: background dose: 2 μg/h, single dose: 2 μg/time, and lockout time: 15 min. The patients will be instructed to press the button on the intravenous analgesia pump (as a remedial analgesic measure) when the numeric rating scale (NRS) scores for pain will reach ≥ 4 points.

### Outcome measures

The investigators will communicate with each patient on preoperative day 1, and will provide guidance regarding the use of PCIA pumps. The NRS will be used to evaluate pain intensity in all cases [19]. In this context, the NRS is a commonly used scoring tool that helps assess the intensity of pain experienced by patients. The intensity is scored on a scale of 0 to 10, where 0 and 10 indicate no pain and the most severe pain, respectively. Patients will be asked to select a number to describe their current level of pain. Usually, scores of 0, 1–3, 4–6, 7–9, and 10 indicate no, mild, moderate, severe, and the most severe pain, respectively.

Efficacy will be measured in terms of the incidence of CPTP at 3 months after surgery; this will be the primary outcome measure of this study. Individuals who meet the diagnostic criteria for CPSP based on the International Association for the Study of Pain (International Classification of Diseases-11) standards will be recruited [4]. Notably, CPSP is defined as pain that persists or increases in intensity for more than 3 months following surgery or tissue injury (including any trauma such as burns). The pain may not be limited to the original surgical or injury site, and may radiate to the entire area innervated by the nerves that supply that region (e.g., deep somatic or visceral tissues). Other potential causes of pain, such as acute or chronic infections, malignant tumors, or pre-existing chronic pain conditions must be excluded before a diagnosis of CPSP can be made. Although the pain is typically neuropathic by nature, and neuropathic pain mechanisms play a significant role in the pathogenesis, the condition should be diagnosed as CPSP and not neuropathic pain. The etiology needs to be identified, and a diagnosis of chronic primary pain should be made if the cause cannot be determined.

The secondary outcomes will include the dosage of propofol and remifentanil used; NRS pain scores at PACU entry, 30 min after PACU entry, and after surgery (at 12 h, 24 h, and 48 h); number of times the PCIA button is pressed within 24 h and 48 h after surgery; the occurrence of post-operative nausea and vomiting, and other adverse reactions including headache, insomnia, itching, respiratory depression, excessive sedation, urinary retention, allergic reactions, liver injury, and kidney injury; post-operative bowel movement and ambulation times; length of hospital stay; surgeon and patient satisfaction; and the incidence of CPTP at 6 and 12 months after surgery.

In the context of post-operative follow-up, the NRS pain scores can be used during telephone interviews conducted at 3, 6, and 12 months after surgery. These interviews will mainly focus on the assessment of any pain experienced by the patient at the surgical site (NRS score > 0).

The impact of pain on patient quality of daily life will also be evaluated, and the degree will be categorized as follows: no, mild, moderate, and severe [20]. The level of intervention required to manage the pain will also be assessed using the following grading system:

Grade A: No intervention required

Grade B: Rest or reduction in activity

Grade C: Self-administered medications

Grade D: Medical help needed from a healthcare professional

A range of efficacy and adverse outcome variables will be selected as observational indicators to comprehensively assess differences between the control and intervention groups. This will help evaluate the efficacy of intravenous paracetamol and mannitol in the management of CPTP. These outcome variables will not only help in presenting the data, but will also reflect the critical impact on patient health and post-operative recovery. The incidence of CPTP at 3 months after the procedure was selected as the primary outcome variable, as this parameter directly reflects the effectiveness of post-operative pain management; this in turn has significant influence on patient quality of life and the recovery process. Adverse outcome variables such as post-operative nausea and vomiting and respiratory depression, among others (which are common post-operative complications) were selected for this trial. Assessment of the incidence of these complications may offer a more comprehensive understanding on the safety and practicality of the intervention measures.

### Adverse events

Patient safety will be closely monitored throughout the study. Adverse events and side effects will be assessed and recorded using a standardized evaluation tool. These events and side effects may include drug allergies, abnormal liver function, and renal dysfunction. Any adverse events or side effects will be promptly reported to the study team and appropriate measures will be taken to ensure patient safety.

An independent safety monitoring committee will be established to oversee and evaluate patient safety throughout the study. This committee will review all reported adverse events and side effects and provide recommendations for any necessary adjustments to the study protocol.

### Auditing

The trial conduct will be regularly audited by an independent team to ensure compliance with the protocol, Good Clinical Practice (GCP) guidelines, and regulatory requirements. Audits will be conducted quarterly, reviewing study documentation and verifying data accuracy and completeness through on-site visits. Any deviations will be documented and addressed promptly. The auditing process will be independent from investigators and the sponsor, reporting directly to the Institutional Review Board (IRB). Audit findings will be used to identify areas for improvement and implement corrective actions to ensure study validity and reliability.

### Study protocol and data collection

Recruitment, intervention, assessment, and participant visits will be performed as per the following schedule:

#### Enrollment

Patients will be enrolled over a period of 12 months, between July 1, 2023 and April 1, 2024. The flow chart of the trial is shown in Figure 1.

**FIGURE I.**
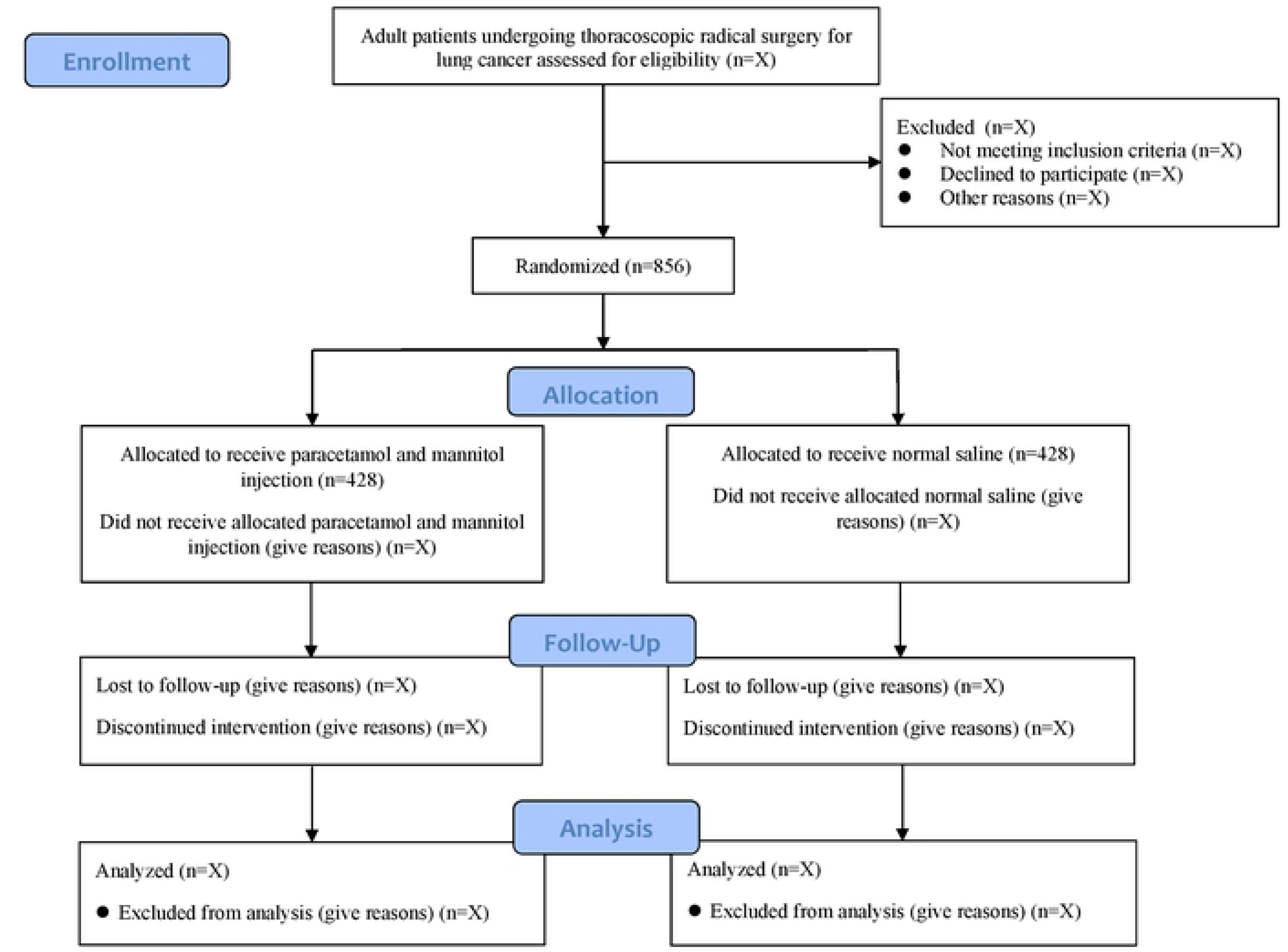
Flow diagram of patients.

#### Interventions

Patients will receive the assigned intervention (intravenous paracetamol and mannitol or normal saline) during thoracoscopic radical surgery for lung cancer.

#### Assessments

Primary outcome assessment for the incidence of CPTP will be performed at 3 months after surgery. Secondary outcome assessments (including dosage of propofol and remifentanil, NRS pain scores, number of times the PCIA button is pressed, occurrence of post-operative nausea and vomiting, respiratory depression, time to first flatus and ambulation, length of hospital stay, surgeon and patient satisfaction, and incidence of CPTP at 6 and 12 months after surgery) will be conducted at specific time points as outlined in the study protocol.

#### Participant visits

Participants will be followed up at 3, 6, and 12 months after surgery to evaluate their quality of daily life. Fig 1 shows the schedule of enrollment, interventions, assessments, and participant visits. This table provides a visual representation of the study timeline and can help ensure adherence to the SPIRIT guidelines.

**Fig 1.**
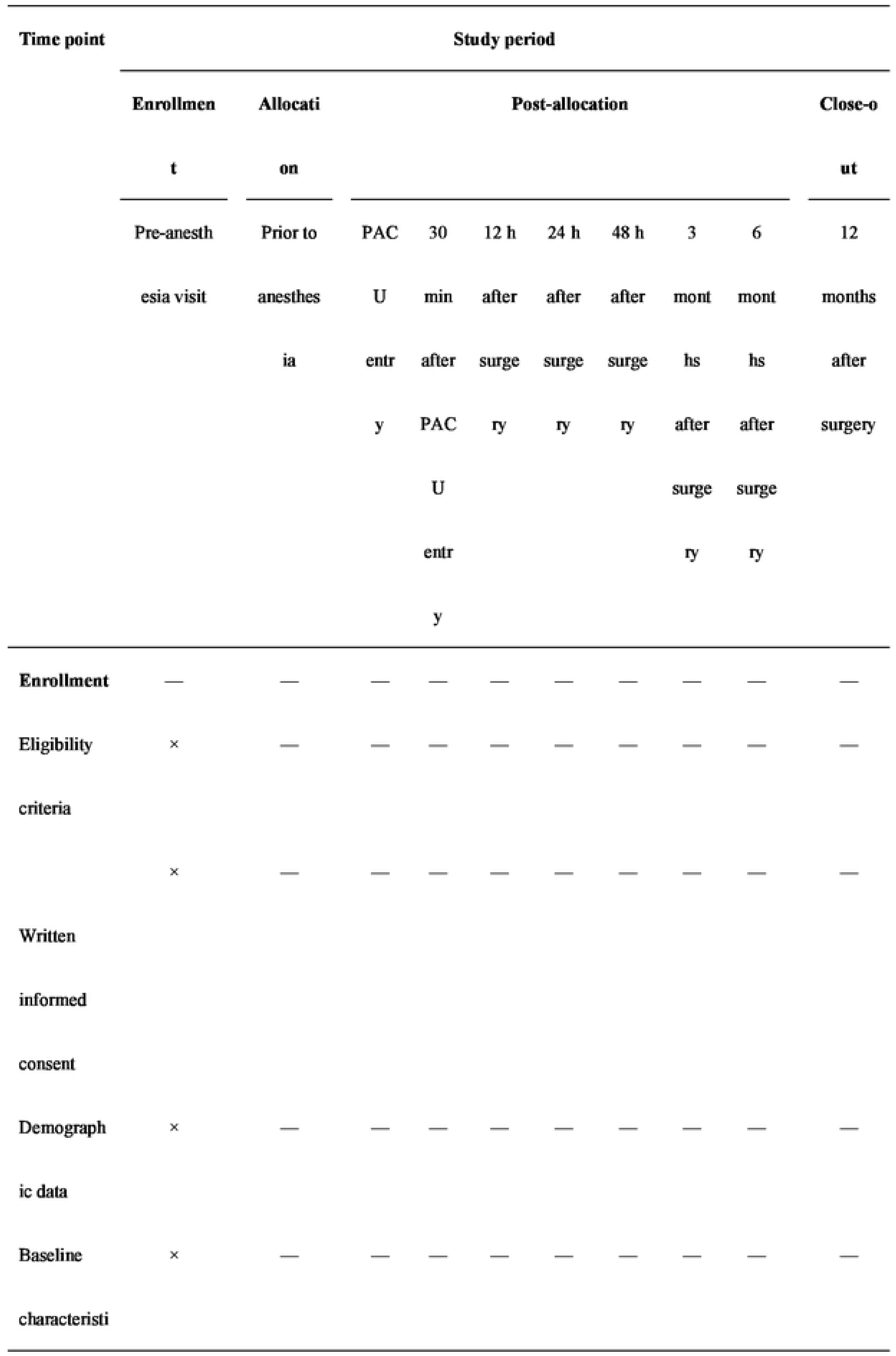
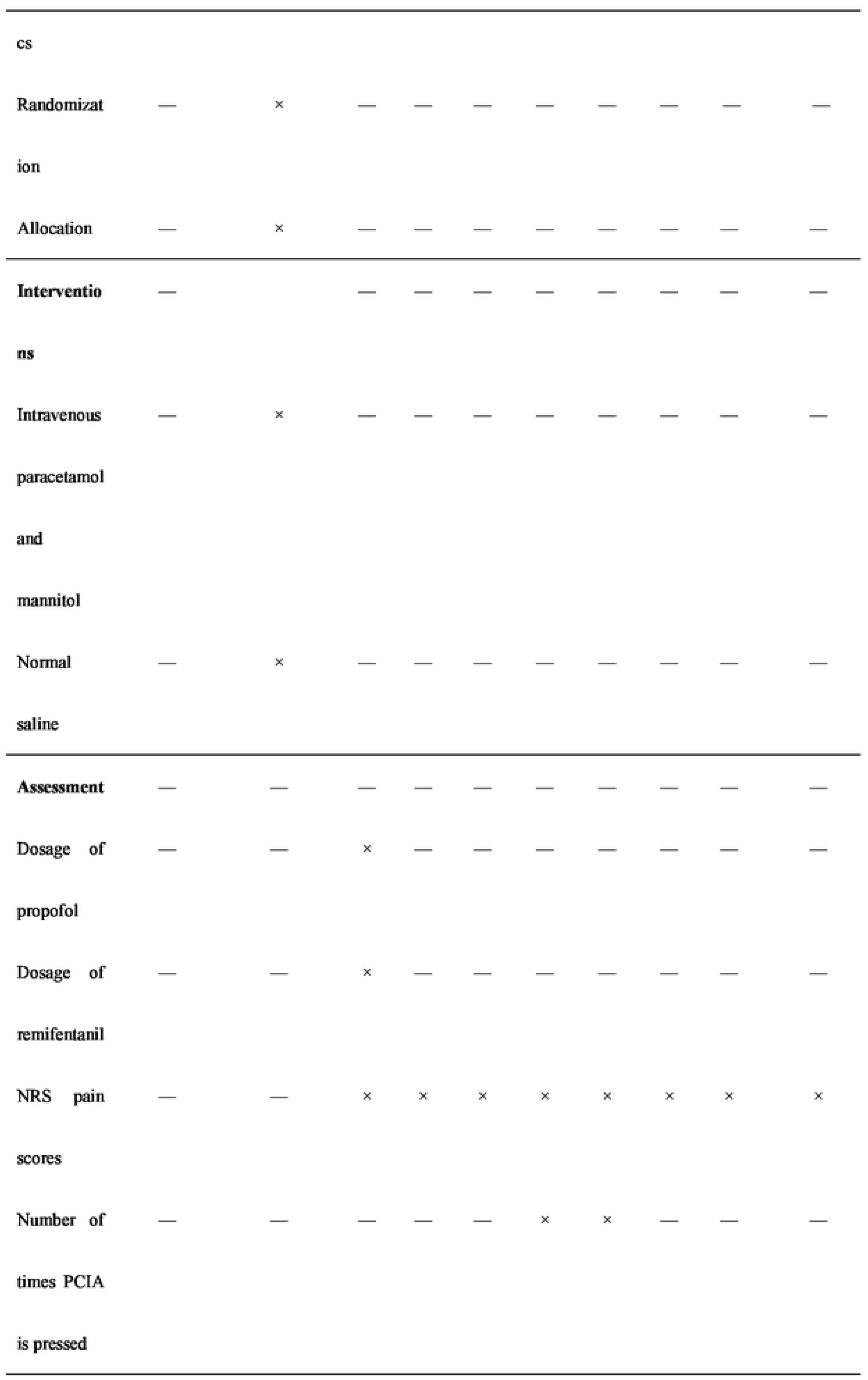
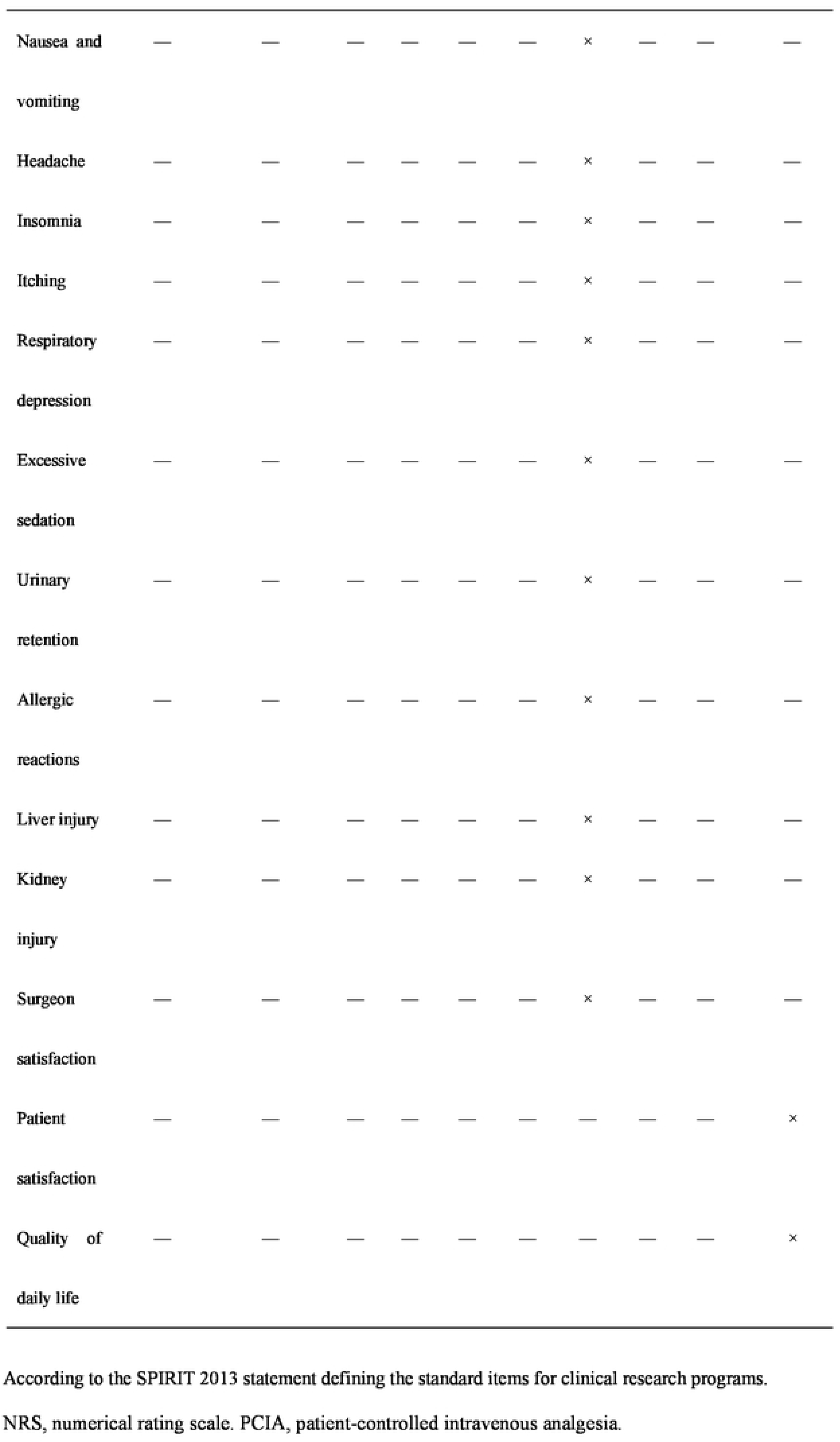
Schedule of patient enrollment, study interventions, and outcomes assessments.

### Data management and monitoring

In this clinical trial, meticulous planning and execution of data entry, coding, security, and storage procedures are essential for safeguarding the integrity and confidentiality of research data. A robust data management framework will be employed for this trial, and it will be overseen by a dedicated team of experts. Key components of this framework will include an electronic data capture (EDC) system for data entry, stringent quality control measures such as double data entry by independent personnel, and rigorous data validation checks to uphold data accuracy. Data management will involve the following specific operational processes:

#### Data entry procedures

Trained research personnel will input data into a secure EDC system, and a dual data entry process will be employed to minimize errors. In cases of discrepancies, data entry operators will perform source data verification to ensure accuracy and consistency.

#### Coding practices

All collected data will be systematically coded using a standardized coding manual to facilitate uniformity and precision in subsequent data analysis. To protect patient confidentiality, any personally identifiable information will be anonymized and replaced with unique study identification numbers.

#### Data security measures

Access to the study database will be strictly controlled and limited to authorized personnel. Data will be stored securely on protected servers with restricted access controls and regular backups to prevent data loss or unauthorized access.

#### Data storage protocols

Electronic data will be housed in a password-protected database with encryption protocols to uphold confidentiality and data security. Physical copies of sensitive study documents will be stored in locked filing cabinets within a secure office environment.

#### Enhancing data quality

The EDC system will be equipped with range checks to validate data values; this will ensure accuracy and completeness. Routine data monitoring and quality control assessments will be conducted to promptly identify and rectify any discrepancies or missing entries.

#### Availability of data management procedures

Detailed data management protocols including the areas of data entry, coding, security, and storage guidelines have been comprehensively outlined in the Data Management Plan document. This document, which delineates specific procedures for handling research data, can be obtained upon request from the principal investigator or the research team.

#### Data collection plan: retention

During the informed consent process, we will provide detailed information to all participants about the study objectives, procedures, potential risks, and benefits. To ensure participant retention and progress monitoring, we will schedule regular follow-up visits or phone calls, reminding them of subsequent appointments. Additionally, we will assign a dedicated study coordinator to address participants’ concerns, answer questions, and provide necessary support throughout the study.

#### Data monitoring: formal committee

The Data Monitoring Committee (DMC) will consist of members with diverse professional backgrounds, including clinical experts, statisticians, and independent ethics experts, ensuring a comprehensive and proficient evaluation of research data. The roles and responsibilities of the DMC will be clearly outlined, emphasizing their duties in monitoring data integrity, accuracy, and safety, evaluating interim analysis results, and providing impartial advice. A structured reporting mechanism will be established to keep the research team and stakeholders informed about the study’s progress and potential issues. The independence of the DMC from the sponsor will be explicitly stated, upholding the credibility and transparency of research data and ensuring the reliability of the findings of this trial.

#### Sample size calculation

The sample size for this study was estimated based on the incidence of CPSP in patients who undergo surgery for lung cancer at 3 months after surgery (the primary indicator for evaluation of efficacy). In this randomized controlled trial, the experimental group will receive intravenous paracetamol and mannitol and the control group will receive normal saline. Studies have shown that the incidence of CPTP ranges from 25% to 57% [3]. It may therefore be assumed that 50% of patients in the control group would develop CPTP at 3 months after surgery. As the pre-trial showed the incidence of CPSP in the trial group to be 40%, this was set as the incidence rate for sample size calculation. Based on a bilateral α value of 0.05 and power (1-β) of 0.8, the sample size ratio between the experimental and control groups was calculated to be 1:1. The sample size was estimated using R, as per the method proposed by Chow et al. [21]. Calculations showed the required sample size in each (intervention and control) group to be 385 cases. On considering a 10% loss-to-follow-up rate, a minimum of 428 cases will be required in either group; a total of 856 cases will therefore be included in this trial.

### Statistical analysis

SPSS 19.0 statistical software (IBM Corporation, Armonk, NY, USA) will be used for all statistical analyses. The two-tailed *P* value will be used to test for statistical significance; *P* < 0.05 will be considered statistically significant. The following statistical analysis methods will be employed:

#### Descriptive analysis

Descriptive statistical analysis will be performed using the demographic variables of the study population including age, sex, and weight. The mean and standard deviation will be reported for continuous variables, and the frequency and percentage will be reported for categorical variables. Normality will initially be tested using the Shapiro– Wilk test. Data conforming to a normal distribution will be presented as means ± standard deviation; other data will be presented as medians with interquartile ranges.

#### Baseline comparisons

The t- or Mann–Whitney U tests (depending on data normality) will be used to compare baseline continuous variables in either group. The chi-square or Fisher’s exact tests will be used to compare intergroup differences between categorical variables.

#### Primary outcome analysis

The primary outcome measure, namely, the incidence of CPTP at 3 months after surgery, will be compared between the test and control groups using appropriate statistical tests (the chi-square or Fisher’s exact tests). Logistic regression may be employed to adjust for potential confounders, as appropriate.

#### Secondary outcome analysis

For continuous variables (such as the dosage of propofol and remifentanil, NRS pain scores, number of times the PCIA button is pressed, time to first flatus and ambulation after surgery, length of hospital stay, and quality of daily life scores), the means or medians will be compared between the two groups using t- or non-parametric tests as appropriate.

Categorical variables including the occurrence of post-operative nausea and vomiting, respiratory depression, surgeon and patient satisfaction, and the incidence of CPTP at 6 and 12 months after surgery will be compared using the chi-square test or logistic regression.

The safety of intravenous paracetamol and mannitol will be evaluated; this will include the incidence of adverse events and complications. The chi-squared or Fisher’s exact tests will be used to determine differences between the two groups.

#### Missing data handling

Missing data will be handled using appropriate methods (such as multiple imputation or sensitivity analysis) to assess the impact of missing data on the results.

#### Subgroup analysis

Subgroup analyses may be conducted based on relevant factors (such as age, sex, type of lung cancer, and preoperative pain scores) to explore potential effect modifiers.

#### Multiplicity adjustment

To control the overall type I error rate, adjustment for multiple comparisons may be considered using methods such as the Bonferroni correction.

#### Interim analysis

Interim analyses will be conducted when 50% and 75% of patients complete the 3-month follow-up, focusing on the primary outcome and safety concerns. An independent Data and Safety Monitoring Board (DSMB) will review these results and provide recommendations. The trial may be stopped early based on statistically significant differences in the incidence of CPTP, safety concerns, or futility, with the final decision made by the Principal Investigator in consultation with the DSMB.

#### Intent-to-treat (ITT) analysis

An ITT analysis will be performed to reduce the impact of bias and increase reliability of the results. Data processing and statistical analysis will be performed in strict adherence with the principles of ITT analysis. All participants will be grouped according to the initial random allocation, regardless of full adherence to the treatment protocol. Appropriate methods will be employed to handle any missing data; this will ensure reliability and objectivity of the analysis. For instance, in cases where a participant has missing follow-up data, suitable methods such as sensitivity analysis will be used based on the initial grouping to assess the impact on the results; appropriate explanations will also be provided during result interpretation.

### Interventions: modifications

All protocol modifications, including changes to eligibility, outcomes, or analyses, will be promptly communicated to investigators, submitted to the Research Ethics Committees (REC)/IRB for approval with detailed rationale and impact assessment, disclosed to participants via updated informed consent, updated in trial registry (Chictr.org.cn.) for transparency, reported in publications, and documented with access for investigators and the trial team to ensure compliance and transparency.

### Interventions: adherance

In this trial, strategies such as education and training, regular monitoring and follow-up, provision of support tools like a calendar, establishment of 24-hour communication channels, and consideration of rewards and incentives will be used to improve patient compliance with the intervention, ultimately ensuring the accuracy and effectiveness of the research results.

### Interventions: concomitant care

During the trial, participants will be allowed to receive routine analgesic therapy, including morphine or other common analgesics, as well as non-interventional pain management methods. They will also be able to receive necessary rehabilitation programs and postoperative care guidance. However, they will be prohibited from taking other experimental analgesic drugs or treatments and from participating in other clinical research projects simultaneously, to ensure the accuracy of the results of this trial.

### Ancillary and post trial care

Participants in this study will receive standard of care for lung cancer and post-thoracotomy pain management, as well as necessary ancillary care throughout the trial and a reasonable post-trial period. Post-trial, they will transition back to regular care providers, with the study team remaining available for guidance and support. A comprehensive compensation plan is in place to protect participants’ rights and welfare, covering medical expenses and other reasonable costs in case of harm directly attributable to trial participation. Adverse events will be promptly reported, investigated, and resolved, with the study sponsor initiating timely and fair compensation when necessary.

### Ethics and dissemination

This trial has been approved by the Ethics Committee of the Henan Provincial Chest Hospital & Chest Hospital of Zhengzhou University on April 29, 2023 (approval number: (2023) EC (04-06)]. The medical ethics approval can be found in Supporting information S3.Written and informed consent will be obtained from all participants prior to their inclusion in this trial. All participants will be provided with detailed information regarding the study objectives, procedures, potential risks and benefits, confidentiality measures, and their right to withdraw from the study at any time. The confidentiality and privacy of participants will be strictly maintained throughout this trial, and it will be conducted in accordance with ethical principles and regulations to ensure that the rights and well-being of all participants are protected.

The results of this trial will be communicated to the participants and subsequently submitted for publication in peer-reviewed journals for wider dissemination. The investigators will actively participate in relevant academic conferences and seminars to share research methodologies and achievements with peers; this will foster academic exchange and collaboration.

## Discussion

The combination of intravenous paracetamol and mannitol characteristically demonstrates rapid onset (peak time of 15 min), high blood drug concentrations (C_max_ of 25–30 μg/mL for a single adult dose of 1000 mg injection), lack of first-pass effect, predictable pharmacokinetics, broad applicability, and efficacy in pain management. Although the traditional view attributed the analgesic action of paracetamol to cyclooxygenase inhibition [22], recent research indicates that the primary mechanism involves the metabolism of N-acylphenol amine (AM404), which subsequently interacts with transient receptor potential vanilloid 1 and cannabinoid 1 receptors in the brain. The paracetamol metabolite (AM404) directly induces analgesia via the transient receptor potential vanilloid 1 receptor at the C-fiber terminals in the spinal dorsal horn [23]. In view of the characteristics and mechanisms of action, the novel combination of intravenous paracetamol and mannitol demonstrates superior analgesic effects and a lower incidence of adverse reactions; this makes it a safe and effective option for pain management. This trial will therefore aim to evaluate the efficacy and safety of the combination in managing CPTP; the results are expected to provide providing valuable insights for future clinical practice.

This trial will focus on effective data collection and accurate interpretation of clinical outcomes, as the effectiveness of data collection is essential for research reliability. It is also essential that the issue of missing data is effectively addressed; transparency will therefore be maintained while reporting the extent of missing data, and robust strategies (such as data imputation and sensitivity analyses) will be employed to reduce bias. Rigorous methodology (such as double data entry and validation checks) will be followed to minimize data entry errors and enhance data quality. The clinical relevance of outcome variations across different cohorts will be elucidated while interpreting clinical outcomes; this will provide insights into patient care implications. For instance, superior pain scores in the intervention group may suggest reduced reliance on analgesics; this can minimize medication-related adverse effects. Our analysis will contextualize the clinical significance of findings; this will enable comparisons with findings in existing literature, and thereby improve comprehension and applicability. As it is crucial to address study limitations (including potential bias related to selection and information), these will be discussed and their impact on outcomes will be evaluated; this will help ensure that the interpretations are comprehensive. Adequate sample size determination is key to robust statistical analyses and reliable conclusions. The rationale behind sample size determination has therefore been discussed; the impact of sample size inadequacy on study outcomes has also been described.

The findings of this study will have significant clinical implications as it aims to evaluate the efficacy of a new combination, namely, intravenous acetaminophen and mannitol, in managing CPTP following surgery for lung cancer. The findings will have the potential to influence clinical practice in several key ways. First, confirmation of the effectiveness of acetaminophen and mannitol in alleviating CPTP could help optimize clinical treatment strategies. This new treatment option may be integrated into post-operative pain management protocols for those undergoing lung cancer surgery, and may thereby enhance patient care. Second, confirmation of superiority of the combination could help reduce the use of propofol and remifentanil for analgesia. This may in turn reduce the frequency of drug-related side effects and complications, and thereby improve post-operative comfort and expedite recovery; this may provide substantial clinical benefits. Finally, evaluation of the impact of the novel combination on quality of life will offer a more comprehensive understanding of the post-operative experience. If intravenous acetaminophen and mannitol are found to improve patient quality of life, the findings could provoke renewed focus on post-operative care practices and enhance overall patient well-being.

## Conclusion

In conclusion, this study protocol outlines a rigorous and comprehensive approach for evaluating the efficacy and safety of intravenous paracetamol and mannitol in managing CPTP after surgery for lung cancer. The study will employ a randomized double-blind controlled design to provide reliable evidence regarding the potential benefits of this treatment approach. The findings will contribute to the existing literature on CPTP management, and may have important implications for improving patient outcomes and quality of life.

## Strengths and limitations

The rigorous design of this trial (single-center, randomized, double-blind, and controlled) will ensure high standards for evaluating drug efficacy. Randomization and blinding will minimize bias, and the control group will allow for direct comparison to accurately assess efficacy. Standardized pain assessment tools will ensure consistency and reliability of outcome measurements. In addition, the single-center design will minimize variability and improve internal validity; however, this may limit generalizability.

The limitations of this trial include potential issues with the sample size, selection bias, generalizability, compliance, and the resource-intensive nature of design and execution.

## Author contributions

JZ and XL will jointly contribute to the design and financial support of this trial, and have also jointly contributed to writing and revision of the manuscript. These two authors have contributed equally to the work and are co-first authors. JG and SL will be responsible for data analysis and will provide relevant research recommendations. WZ and GX will be responsible for the specific implementation of the research program. YG will assist in data collection. HX will be responsible for the design and quality control of the study, provide relevant research suggestions, and participate in manuscript revision. All authors will contribute to the completion of the study and the writing of the manuscript. All authors agree to be accountable for the content of the work.

## Data Availability

Deidentified research data will be made publicly available when the study is completed and published.

N/A.

## Acknowledgements

The authors would like to thank Xia Gao and Xiao-bo Wang for their contributions to research quality assurance and data validation.

## Supporting information

S1 Checklist. SPIRIT checklist.(DOC)

S1 Protocol. Study protocol.(DOCX)

S2 The template for the informed consent form

S3 Medical Ethics Approval

## Patient and public involvement

Patients and/or the public were not involved in the design, or conduct, or reporting, or dissemination plans of this trial.

## Patient consent for publication

Not required.

## Data availability

During the current study, no datasets were generated or analyzed. All pertinent data from this study will be accessible upon its completion.

## References

1. Lopes A, Seligman Menezes M, Antonio Moreira de Barros G. Chronic postoperative pain: ubiquitous and scarcely appraised: narrative review. Braz J Anesthesiol. 2021;71: 649–655.

2. Marshall K, McLaughlin K. Pain management in thoracic surgery. Thorac Surg Clin. 2020;30: 339–346.

3. Gupta R, Van De Ven T, Pyati S. Post-thoracotomy pain: current strategies for prevention and treatment. Drugs. 2020;80: 1677–1684.

4. Schug SA, Lavand’homme P, Barke A, Korwisi B, Rief W, Treede RD. The IASP classification of chronic pain for ICD-11: chronic postsurgical or posttraumatic pain. Pain. 2019;160: 45–52.

5. Reznik SI. Post-thoracotomy pain and nerve protection: back to the drawing board? J Thorac Cardiovasc Surg. 2019;157: 376–377.

6. Wildgaard K, Ravn J, Kehlet H. Chronic post-thoracotomy pain: a critical review of pathogenic mechanisms and strategies for prevention. Eur J Cardiothorac Surg. 2009;36: 170–180.

7. Fortier S, Hanna HA, Bernard A, Girard C. Comparison between systemic analgesia, continuous wound catheter analgesia and continuous thoracic paravertebral block: a randomised, controlled trial of postthoracotomy pain management. Eur J Anaesthesiol. 2012;29: 524–530.

8. Mijatovic D, Bhalla T, Farid I. Post-thoracotomy analgesia. Saudi J Anaesth. 2021;15: 341–347.

9. Coppens S, Eochagain AN, Hoogma DF, Dewinter G. Stranger things: the erector spinae block, extra sensory perception, or paranormal block by proxy? APS. 2023;1:12. doi: 10.1007/s44254-023-00007-5.

10. Pasero C, Stannard D. The role of intravenous acetaminophen in acute pain management: a case-illustrated review. Pain Manag Nurs. 2012;13: 107–124.

11. American Society of Anesthesiologists Task Force on Acute Pain Management. Practice guidelines for acute pain management in the perioperative setting: an updated report by the American society of anesthesiologists task force on acute pain management. Anesthesiology. 2012;116: 248–273.

12. Singla NK, Hale ME, Davis JC, Bekker A, Gimbel J, Jahr J, et al. IV acetaminophen: efficacy of a single dose for postoperative pain after hip arthroplasty: subset data analysis of 2 unpublished randomized clinical trials. Am J Ther. 2015;22: 2–10.

13. Nishimoto RN. OFIRMEV: an old drug becomes new again. Anesth Prog. 2014;61: 99–102.

14. Ghaffarpasand F, Dadgostar E, Ilami G, Shoaee F, Niakan A, Aghabaklou S, et al. Intravenous acetaminophen (paracetamol) for postcraniotomy pain: systematic review and meta-analysis of randomized controlled trials. World Neurosurg. 2020; 134: 569–576.

15. Nonaka T, Hara M, Miyamoto C, Sugita M, Yamamoto T. Comparison of the analgesic effect of intravenous acetaminophen with that of flurbiprofen axetil on post-breast surgery pain: a randomized controlled trial. J Anesth. 2016;30: 405–409.

16. Dastan F, Langari ZM, Salamzadeh J, Khalili A, Aqajani S, Jahangirifard A. A comparative study of the analgesic effects of intravenous ketorolac, paracetamol, and morphine in patients undergoing video-assisted thoracoscopic surgery: a double-blind, active-controlled, randomized clinical trial. Ann Card Anaesth. 2020;23: 177–182.

17. Sharma CV, Long JH, Shah S, Rahman J, Perrett D, Ayoub SS, et al. First evidence of the conversion of paracetamol to AM404 in human cerebrospinal fluid. J Pain Res. 2017;10: 2703–2709.

18. Chan A-W, Tetzlaff JM, Gøtzsche PC, Altman DG, Mann H, Berlin J, et al. SPIRIT 2013 explanation and elaboration: guidance for protocols of clinical trials. BMJ. 2013;346: e7586.

19. Karcioglu O, Topacoglu H, Dikme O, Dikme O. A systematic review of the pain scales in adults: which to use? Am J Emerg Med. 2018;36: 707–714.

20. Kumar SP. Utilization of brief pain inventory as an assessment tool for pain in patients with cancer: a focused review. Indian J Palliat Care. 2011;17: 108–115.

21. Chow SC, Shao J, Wang H. Sample size calculation in clinical research. 2nd Ed. New York: Chapman & Hall/CRC Press; 2007.

22. Ayoub SS. Paracetamol (acetaminophen): A familiar drug with an unexplained mechanism of action. Temperature (Austin). 2021;8: 351–371.

23. Ohashi N, Kohno T. Analgesic effect of acetaminophen: a review of known and novel mechanisms of action. Front Pharmacol. 2020;11: 580289.

